# Serologic Evidence of Crimean-Congo Hemorrhagic Fever Virus Exposure among Livestock and Farmers in the Democratic Republic of the Congo

**DOI:** 10.1101/2025.08.18.25333611

**Authors:** Megan Halbrook, Boniface Lombe, Sydney Merritt, Emmanuel Hasivirwe Vakaniaki, Daddy Kanonge, Charlotte Tshingula, Yannick Munyeku-Bazitama, Masahiro Kajihara, Sheila Makiala-Mandanda, Ryan Harrigan, Patrick Mukadi-Kakoni, Nicole A. Hoff, Steve Ahuka Mundeke, Ayato Takada, Augustin Twabela, Placide Mbala-Kingebeni, Lisa Hensley, Anne W. Rimoin

## Abstract

Crimean-Congo Hemorrhagic Fever (CCHF) is a potential high-threat zoonotic disease caused by the Crimean-Congo hemorrhagic fever virus (CCHFV). Transmission of CCHFV occurs primarily through bites of infected *Hyalomma* ticks or via direct contact with the blood or tissues of infected animals or humans. This study presents a cross-sectional assessment of CCHFV seroprevalence and risk factors associated with occupational and environmental exposures among cattle and swine agricultural workers.

Nine provinces across the Democratic Republic of the Congo (DRC) were selected and collection took place from June 2023 to July 2024. Five herds per species in each province were randomly visited, and at each facility or herd, up to 20 animals were chosen for serum sampling and attached tick collection. In five provinces, farm workers present on the day of collection were enrolled. Detection of anti-CCHFV Immunoglobulin G (IgG) antibodies was assessed via an in-house nucleoprotein-based enzyme-linked immunosorbent assay (ELISA).

Among the 1,118 cattle surveyed across nine provinces 57.0% (95%CI: 54.1-59.9%) were seroreactive. Cattle from two provinces in the southeast, Tanganyika and Lualaba, had 94.6% (95%CI: 89.9-99.2%) and 90.7% (95%CI: 84.9-96.5%) reactivity, respectively.

Among the 1,020 swine surveyed 13.4% (95%CI: 11.1-15.2%) were seroreactive. Among the 180 agricultural workers surveyed, 12.8% (95%CI: 7.9-17.6%) (23) were seroreactive for CCHF antibodies.

This serologic survey indicated that CCHFV is circulating in the DRC and the southeast provinces are particularly at risk for spillover and morbidity among humans. Though no human cases have been reported since 2008, surveillance for CCHF should be considered among veterinary professional and healthcare workers.

## Introduction

Crimean-Congo Hemorrhagic Fever (CCHF) is a potential high-threat zoonotic disease caused by the Crimean-Congo hemorrhagic fever virus (CCHFV), a tick-borne virus belonging to the Nairoviridae family, genus *Orthonairovirus*^1,2^. The disease is characterized by the abrupt onset of high fever, headache, myalgia, gastrointestinal symptoms, and in severe cases, hemorrhagic manifestations, multi-organ failure, and death. With a case fatality rate estimated between 10% and 40%, CCHF is among the most virulent of hemorrhagic fevers affecting humans^3,4^. The World Health Organization (WHO) has designated CCHF as a priority pathogen and a target of global epidemic preparedness initiatives due to its growing geographical distribution, severe clinical outcomes in humans, potential for nosocomial outbreaks, and lack of mitigation strategies^5,6^.

Transmission of CCHFV occurs primarily through bites of infected *Hyalomma* ticks or via direct contact with the blood or tissues of infected animals or humans^7^. Livestock, such as cattle, goats, and sheep, can be infected and while livestock mainly experience asymptomatic infection, they are key species in the transmission cycle of the virus to other hosts and humans^8^. Populations at highest risk for disease transmission include farmers, herders, veterinarians, abattoir workers, and butchers, who may encounter infectious materials from animals or humans, or tick infestations through routine animal husbandry or slaughter activities^9,10^. This occupational risk is exacerbated in low- and middle-income countries, where protective equipment, training, and veterinary oversight are often limited.

The Democratic Republic of the Congo (DRC), the second-largest country in Africa in terms of land area, encompassing vast ecological zones that are favorable for maintaining both tick populations and supporting livestock production. Agriculture and animal husbandry comprise a significant portion of the country’s informal economy and livelihood base, particularly in rural areas^11^. The DRC has a fragile public health infrastructure, limited veterinary surveillance, and lack of routine diagnostic capacity that has hampered efforts to monitor or quantify zoonotic diseases such as CCHF. Since the virus was first described and documented in Crimea in the 1940s and first virus isolation in DRC in the 1950s, epidemiological data regarding the scope of CCHF disease risk in central Africa remain sparse and fragmented^12^. To date, there has been no comprehensive One Health assessment of CCHFV prevalence among Congolese livestock and persons who work in close contact with livestock.

This study presents a cross-sectional assessment of CCHFV seroprevalence and risk factors associated with occupational and environmental exposures among livestock (cattle and swine), persons with occupational exposure to livestock including farmers, herders, and abattoir workers. This is among the first integrated One Health studies of CCHFV prevalence in the DRC which links animal and farmer serosurveillance with farm management practices and vector distribution. The results from this study could be used to inform both national policy and international preparedness frameworks. Moreover, they may help to close a critical knowledge gap in an under-studied region of sub-Saharan Africa and to illustrate how strategic global health engagement aligns with national security objectives. The proactive detection of zoonotic viruses, such as CCHFV, at their source contributes not only to the protection of vulnerable populations in endemic areas but also to safeguarding the health and stability of nations far beyond their borders.

## Methods

Nine provinces across the DRC were selected based on livestock density reports provided by the Central Veterinary Laboratory in Kinshasa and logistical feasibility: Tanganyika, Lualaba, Kasai, Tshopo, Kasai Oriental. Maniema, Kinshasa, Equateur, and Sud Ubangi. Collection took place from June 2023 to July 2024. Five herds per species in each province were randomly visited, and at each facility or herd, up to 20 animals were chosen for serum sampling and attached tick collection. A brief health survey was conducted for each animal which assessed body condition score, tick load, the presence of lesions, and current illness state, if any. A facility survey was administered to the farm owner or manager to assess the facility management practices that may impact CCHF transmission such as grazing habits, availability of medicines, history of infectious disease among the herd, and others. Up to 20 ticks found on each animal were collected. The dorsal and ventral sides of collected ticks were imaged and speciated using deep-learning algorithms via the IDX machine (Vectech, Baltimore, MD, USA) and confirmed by trained lab staff. In five provinces (Kasai Oriental, Kinshasa, Lualaba, Maniema, Sud Ubangi), all personnel present at the facility were invited to enroll in the study and a serum sample and survey on demographics and occupational and personal risks was collected.

Detection of anti-CCHFV Immunoglobulin G (IgG) antibodies was assessed via an in-house enzyme-linked immunosorbent assay (ELISA) that has previously been described^13^. All sera were heat-inactivated at 56 °C for 30 minutes before testing. The assay involved coating plates with purified CCHFV NP antigen and blocking with 3% skim milk in phosphate-buffered saline (PBS). Serum samples, diluted 1:100 in PBS with 1% skim milk, were then added. Bound antibodies were detected using a horseradish peroxidase–conjugated protein A/G followed by the addition of 3,3’,5,5’-tetramethylbenzidine (TMB) substrate. The reaction was stopped with 1 N phosphoric acid, and absorbance was measured at 450 nm. The assay was validated using a panel of controls, including mouse anti-NP monoclonal antibodies, CCHFV IgG-positive human sera, and naïve human sera. To assess potential cross-reactivity, CCHFV IgG-positive animal samples were also tested for anti-Nairobi sheep disease virus (NSDV) NP IgG. All samples were tested in duplicate, and the mean optical density (OD) values were used for analysis. The assay cut-off and interpretation criteria were determined statistically based on the established protocol^13^.

The relationship between seroreactivity and survey data was assessed via chi-square analysis using an alpha of 0.05 in SAS 9.6 (Cary, NC).

Ethics approvals for this work was provided by Institutional Review Boards of the University of California, Los Angeles (IRB#23-000676) and the Kinshasa School of Public Health, University of Kinshasa, DRC (ESP/CE/161/2024). Animal research ethical review was provided by of the University of California, Los Angeles (ARC#2023-009).

## Results

Among the 1,020 swine surveyed 13.4% (95%CI: 11.1-15.2%) were seroreactive. Swine from Kasai province had 42.1% (95%CI: 32.2-52.0%) seroreactivity, the highest rate of seroreactivity among swine (Figure 1). Attached ticks were found on 592 (52.9%) of all cattle and 4 (0.4%) of swine at the time of collection; seroreactivity was not associated with the presence of ticks (Table 1). Among the collected ticks, 16 *Hyalomma rufipes* were identified on 9 cattle from four provinces: Kinshasa (6), Sud-Ubangi (1), Kasai (1), and Tanganyika (8). Among these 9 cattle, 6 were seroreactive for CCHF. Very few (<1%) cattle and swine were experiencing illness or exhibiting symptoms at the time of data collection.

**Table 1.** Livestock Seroreactivity.

|  | Cattle |  |  |  |  | Swine |  |  |  |  |
| --- | --- | --- | --- | --- | --- | --- | --- | --- | --- | --- |
|  | All |  | Seroreactive |  |  | All |  | Seroreactive |  |  |
|  | N=1118 |  | n= 637 (57.0%) |  |  | N=1020 |  | n= 134 (13.4%) |  |  |
|  | N | ColPct | N | ColPct | RowPct | N | ColPct | N | ColPct | RowPct |
| **Province** |  |  |  |  |  |  |  |  |  |  |
| Tanganyika | 92 | 8.2 | 87 | 13.7 | 94.6 | 96 | 9.4 | 6 | 4.5 | 6.3 |
| Lualaba | 97 | 8.7 | 88 | 13.8 | 90.7 | 97 | 9.5 | 12 | 9.0 | 12.4 |
| Kasai | 100 | 8.9 | 77 | 12.1 | 77.0 | 95 | 9.3 | 40 | 29.9 | 42.1 |
| Tshopo | 99 | 8.9 | 70 | 11.0 | 70.7 | 92 | 9.0 | 18 | 13.4 | 19.6 |
| Kasai Oriental | 98 | 8.8 | 64 | 10.1 | 65.3 | 100 | 9.8 | 7 | 5.2 | 7.0 |
| Maniema | 79 | 7.1 | 48 | 7.5 | 60.8 | 99 | 9.7 | 18 | 13.4 | 18.2 |
| Kinshasa | 355 | 31.8 | 145 | 22.8 | 40.9 | 282 | 27.6 | 21 | 15.7 | 7.5 |
| Equateur | 101 | 9.0 | 30 | 4.7 | 29.7 | 98 | 9.6 | 3 | 2.2 | 3.1 |
| Sud Ubangi | 97 | 8.7 | 28 | 4.4 | 28.9 | 61 | 6.0 | 9 | 6.7 | 14.8 |
| **Sex** |  |  |  |  |  |  |  |  |  |  |
| Female | 749 | 67.0 | 467 | 73.3 | 62.4 | 665 | 65.2 | 103 | 76.9 | 15.5 |
| Male | 369 | 33.0 | 170 | 26.7 | 46.1 | 355 | 34.8 | 31 | 23.1 | 8.7 |
| **Age** |  |  |  |  |  |  |  |  |  |  |
| Infant | 117 | 10.5 | 45 | 7.1 | 38.5 | 31 | 3.0 | 4 | 3.0 | 12.9 |
| Juvenile | 240 | 21.5 | 119 | 18.7 | 49.6 | 426 | 41.8 | 43 | 32.1 | 10.1 |
| Adult | 745 | 66.6 | 465 | 73.0 | 62.4 | 561 | 55.0 | 86 | 64.2 | 15.3 |
| Senior | 15 | 1.3 | 8 | 1.3 | 53.3 | 1 | 0.1 | 1 | 0.8 | 100.0 |
| Geriatric | 1 | 0.1 | 0 | 0.0 | 0.0 | 1 | 0.1 | 0 | 0.0 | 0.0 |
| **Body Condition Score** |  |  |  |  |  |  |  |  |  |  |
| 2 | 16 | 1.4 | 5 | 0.8 | 31.3 | 73 | 7.2 | 5 | 3.7 | 6.9 |
| 3 | 298 | 26.7 | 148 | 23.2 | 49.7 | 514 | 50.4 | 58 | 43.3 | 11.3 |
| 4 | 612 | 54.7 | 393 | 61.7 | 64.2 | 381 | 37.4 | 60 | 44.8 | 15.8 |
| 5 | 154 | 13.8 | 73 | 11.5 | 47.4 | 48 | 4.7 | 9 | 6.7 | 18.8 |
| 6 | 38 | 3.4 | 18 | 2.8 | 47.4 | 4 | 0.4 | 2 | 1.5 | 50.0 |
| **Lesions** |  |  |  |  |  |  |  |  |  |  |
| No | 1073 | 96.0 | 613 | 96.2 | 57.1 | 975 | 95.6 | 127 | 94.8 | 13.0 |
| Yes | 45 | 4.0 | 24 | 3.8 | 53.3 | 45 | 4.4 | 7 | 5.2 | 15.6 |
| **Ticks** |  |  |  |  |  |  |  |  |  |  |
| No | 526 | 47.1 | 298 | 46.8 | 56.7 | 1016 | 99.6 | 134 | 100.0 | 13.2 |
| Yes | 592 | 53.0 | 339 | 53.2 | 57.3 | 4 | 0.4 | 0 | 0.0 | 0.0 |

**Figure 1.**
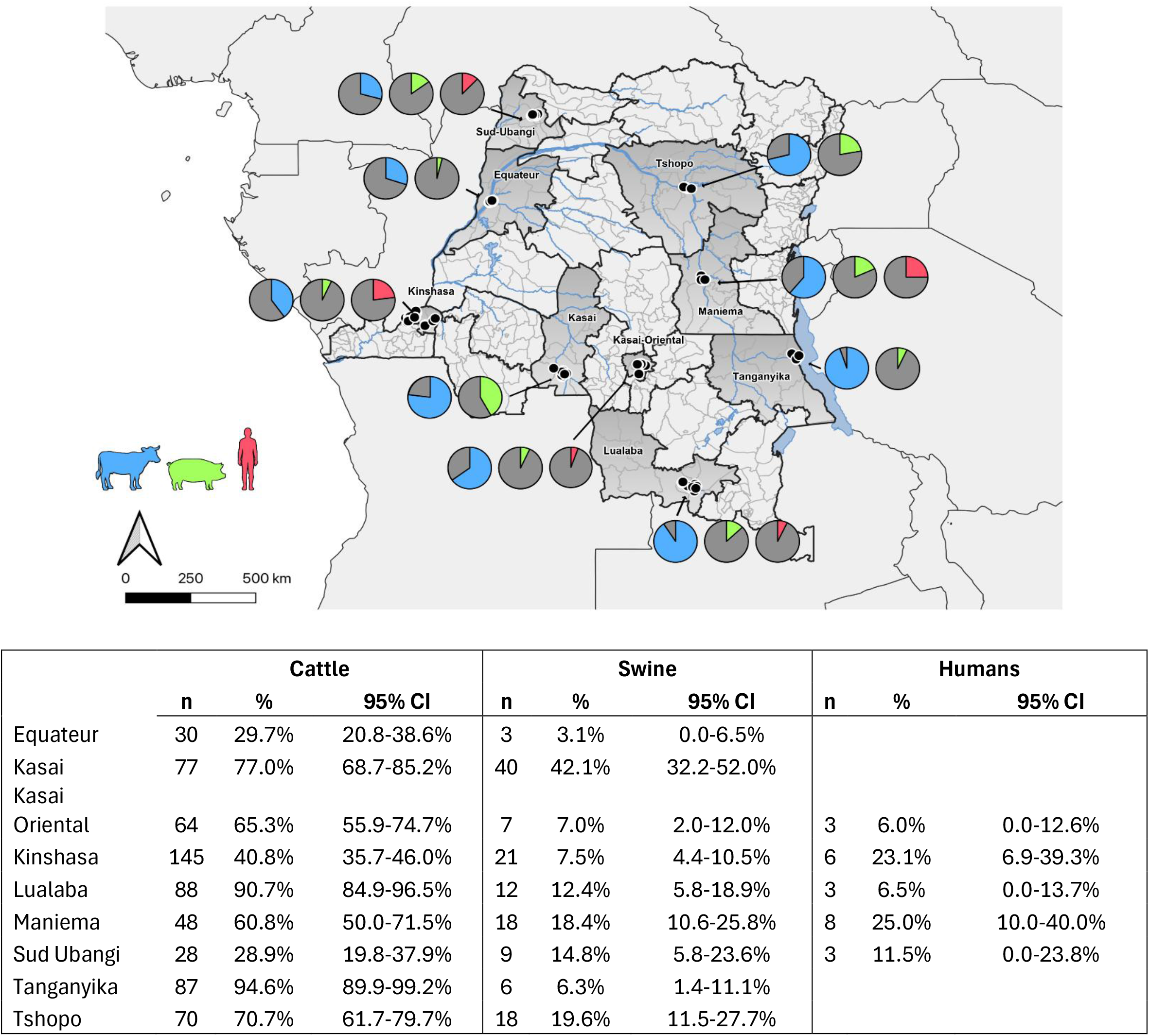
CCHF seroreactivity among cattle, swine, and agriculture workers in the Democratic Republic of the Congo

Of the 180 agricultural workers surveyed, 12.8% (95%CI: 7.9-17.6%) (23) were seroreactive for CCHF antibodies. Maniema and Kinshasa provinces had the greatest number of seroreactive individuals, 8 (25%, 95%CI: 10.0-40.0%) and 6 (26.1%, 95%CI: 6.9-39.3%), respectively. Among all participants, 65.6% reported that they had contact with a sick animal in the past year. While there was a slightly greater proportion of seroreactivity among those who had contact compared with those who did not (15.2% vs 8.5%), the relationship was not statistically significant (Chi-sq p= 0.2060). The use of personal protective equipment (PPE) was limited as 80% reported that they rarely or never wear PPE. 55% of participants reported that they live on the farm property. Across the cohort, 16.1% reported that they never wash their hands during their workday, 35.6% reported handwashing once or twice daily. Outside of work, 86.1% of agricultural workers reported that they entered the forest, 34.4% reported hunting wild animals, and 53.3% reported eating bushmeat (Table 2).

**Table 2.** Farmer Demographics by Seroreactivity.

|  |  | All<br>N= 180 |  | Negative<br>n = 157 (87.2) |  |  | Seroreactive<br>n= 23 (12.8) |  |  |
| --- | --- | --- | --- | --- | --- | --- | --- | --- | --- |
|  |  | N | ColPct | N | ColPct | RowPct | N | ColPct | RowPct |
| **Province** |  |  |  |  |  |  |  |  |  |
|  | Kasai Oriental | 50 | 27.78 | 47 | 29.94 | 94 | 3 | 13.04 | 6 |
|  | Kinshasa | 26 | 14.4 | 20 | 12.7 | 76.9 | 6 | 26.09 | 23.1 |
|  | Lualaba | 46 | 25.56 | 43 | 27.39 | 93.48 | 3 | 13.04 | 6.52 |
|  | Maniema | 32 | 17.78 | 24 | 15.29 | 75 | 8 | 34.78 | 25 |
|  | Sud Ubangi | 26 | 14.44 | 23 | 14.65 | 88.46 | 3 | 13.04 | 11.54 |
| **Site Type** |  |  |  |  |  |  |  |  |  |
|  | Commerical farm | 132 | 73.33 | 115 | 73.25 | 87.12 | 17 | 73.91 | 12.88 |
|  | Village farm | 33 | 18.33 | 29 | 18.47 | 87.88 | 4 | 17.39 | 12.12 |
|  | Abbatoir | 15 | 8.33 | 13 | 8.28 | 86.67 | 2 | 8.7 | 13.33 |
| **Age** |  |  |  |  |  |  |  |  |  |
|  | 18-24 | 40 | 22.22 | 37 | 23.57 | 92.5 | 3 | 13.04 | 7.5 |
|  | 25-34 | 44 | 24.44 | 41 | 26.11 | 93.18 | 3 | 13.04 | 6.82 |
|  | 35-49 | 52 | 28.89 | 47 | 29.94 | 90.38 | 5 | 21.74 | 9.62 |
|  | 50-75 | 44 | 24.44 | 32 | 20.38 | 72.73 | 12 | 52.17 | 27.27 |
| **Sex** |  |  |  |  |  |  |  |  |  |
|  | Female | 36 | 20 | 33 | 21.02 | 91.67 | 3 | 13.04 | 8.33 |
|  | Male | 144 | 80 | 124 | 78.98 | 86.11 | 20 | 86.96 | 13.89 |
| **Primary Residence at Farm** |  |  |  |  |  |  |  |  |  |
|  | No | 81 | 45 | 68 | 43.32 | 134.81 | 13 | 56.52 | 65.19 |
|  | Yes | 99 | 55 | 89 | 56.69 | 89.9 | 10 | 43.48 | 10.1 |
| **Contact with sick animals in last 12 months** |  |  |  |  |  |  |  |  |  |
|  | No | 59 | 32.78 | 54 | 35.06 | 91.53 | 5 | 21.74 | 8.47 |
|  | Yes | 118 | 65.56 | 100 | 64.94 | 84.75 | 18 | 78.26 | 15.25 |
| **PPE Frequency** |  |  |  |  |  |  |  |  |  |
|  | Never | 124 | 68.89 | 109 | 69.43 | 87.9 | 15 | 65.22 | 12.1 |
|  | Rarely | 20 | 11.11 | 16 | 10.19 | 80 | 4 | 17.39 | 20 |
|  | Sometimes | 11 | 6.11 | 10 | 6.37 | 90.91 | 1 | 4.35 | 9.09 |
|  | Often | 6 | 3.33 | 4 | 2.55 | 66.67 | 2 | 8.7 | 33.33 |
|  | Always | 19 | 10.56 | 18 | 11.46 | 94.74 | 1 | 4.35 | 5.26 |
| **Handwashing** |  |  |  |  |  |  |  |  |  |
|  | Never | 29 | 16.11 | 27 | 17.2 | 93.1 | 2 | 8.7 | 6.9 |
|  | 1-2 times per day | 64 | 35.56 | 58 | 36.94 | 90.63 | 6 | 26.09 | 9.38 |
|  | Few times a day (3-5) | 26 | 14.44 | 19 | 12.1 | 73.08 | 7 | 30.43 | 26.92 |
|  | Many times a day | 61 | 33.89 | 53 | 33.76 | 86.89 | 8 | 34.78 | 13.11 |
| **Have you ever done the following activites?** |  |  |  |  |  |  |  |  |  |
|  | Enter a cave or mine | 27 | 15 | 24 | 15.29 | 88.89 | 3 | 13.04 | 11.11 |
|  | Visit the forest | 155 | 86.11 | 135 | 85.99 | 87.1 | 20 | 86.96 | 12.9 |
|  | Hunt wild animals | 62 | 34.44 | 54 | 34.39 | 87.1 | 8 | 34.78 | 12.9 |
|  | Slaughter/ butcher wild animals | 80 | 44.44 | 71 | 45.22 | 88.75 | 9 | 39.13 | 11.25 |
|  | Prepared wild animals to eat | 96 | 53.33 | 82 | 52.23 | 85.42 | 14 | 60.87 | 14.58 |

Of the 56 cattle farms visited, all but one farm in Kinshasa province had at least one seroreactive animal. On average, 59.5% of cattle tested at each facility were seroreactive to CCHFV; at seven farms, three in Lualaba province and four in Tanganyika, all animals tested were seroreactive. Among the 65 swine farms visited, 56.9% (37) had at least one seroreactive animal on the day of sampling. On average, we observed 21.5% seroreactivity among selected swine among farms that had at least one seroreactive animal.

All but three (94.5%) cattle farmers reported that they allow their cattle to graze and 87.5% reported that they had an issue with ticks within their herd. Among swine herds, 16.9% allow grazing and 3 farms (4.6%) reported tick problems. During collection it was observed that pigs were often kept in small, wet enclosures unsuitable for ticks compared with grassier and more open areas where cattle were kept. For both cattle and swine, the use of vaccines was less common, 19.6% and 21.5% of farms, respectively; however the use of medicines was very common—73.2% of cattle farms and 67.7% of swine farms regularly used and had access to medicines. When asked about nose-to-nose contact with other species, rats (37.5%), mice (28.6%), and poultry (28.6%) were most common among cattle herds. Among swine herds, 55.4% of farmers reported nose-to-nose contact with rats and mice. Swine farms with at least one seroreactive animal reported much higher rates of contact with rodents compared to those that did not— for rats: 66.7% vs 33.3%, Chi-sq p = 0.0314; for mice: 69.4% vs 30.6%, Chi sq p = 0.0083 (Table 3).

**Table 3.** Farm Management Practices.

|  | Cattle Farms (N= 56) |  | Swine Farms (N= 65) |  |
| --- | --- | --- | --- | --- |
|  | N | % | N | ColPctN |
| **Herd size** |  |  |  |  |
| one-10 | 3 | 5.36 | 7 | 10.77 |
| ten-25 | 10 | 17.86 | 39 | 60 |
| 25-50 | 21 | 37.5 | 13 | 20 |
| 50-100 | 13 | 23.21 | 5 | 7.69 |
| 100-250 | 8 | 14.29 | 1 | 1.54 |
| **History of reporting animal diseases** |  |  |  |  |
| No | 37 | 66.07 | 48 | 73.85 |
| Yes | 19 | 33.93 | 17 | 26.15 |
| **Do you allow your animals to graze?** |  |  |  |  |
| No | 3 | 5.36 | 54 | 83.08 |
| Yes | 53 | 94.64 | 11 | 16.92 |
| **Familiarity with common livestock diseases\*** |  |  |  |  |
| *missing* | 20 | 35.71 | 28 | 43.08 |
| Never heard of it | 6 | 10.71 | 12 | 18.46 |
| Recognized some from the list | 7 | 12.5 | 4 | 6.15 |
| I know some basics | 9 | 16.07 | 13 | 20 |
| I am well informed | 14 | 25 | 8 | 12.31 |
| **Are there any tick problems in your herd?** |  |  |  |  |
| No | 7 | 12.5 | 62 | 95.38 |
| Yes | 49 | 87.5 | 3 | 4.62 |
| **Do you routinely vaccinate your herd?** |  |  |  |  |
| No | 44 | 78.57 | 51 | 78.46 |
| Yes | 11 | 19.64 | 14 | 21.54 |
| **Do you regularly use medicines?** |  |  |  |  |
| No | 15 | 26.79 | 21 | 32.31 |
| Yes | 41 | 73.21 | 44 | 67.69 |
| **Nose-to-nose contact in the last 12 months** |  |  |  |  |
| Bat | 8 | 14.29 | 8 | 12.31 |
| Rat | 21 | 37.5 | 36 | 55.38 |
| Mouse | 16 | 28.57 | 36 | 55.38 |
| Dog | 11 | 19.64 | 13 | 20 |
| Cat | 4 | 7.14 | 12 | 18.46 |
| Poultry | 16 | 28.57 | 14 | 21.54 |
| Monkey | 0 | 0 | 0 | 0 |

\*Cattle diseases listed: Johne's disease, viral diarrhea, brucellosis, tuberculosis, mastitis, mycoplasma, pneumonia, babesiosis, anaplasmosis, foot and mouth disease, anthrax, Contagious bovine pleuropneumonia, rift valley fever, bluetongue, otitis

## Discussion

This survey represents the largest integrated One Health CCHFV serologic survey in the DRC to date. In this paper, we focus on a defined cohort of agricultural workers with direct livestock contact, paired with concurrent sampling of cattle and swine—the first reported CCHFV seroprevalence data for swine in the DRC. In addition to serology, we collected and identified attached ticks, enabling integration of vector ecology with host infection status.

Detailed farm management and biosecurity practices, along with participant behavioral data (PPE use, hygiene practices, wildlife contact), allowed us to identify modifiable on-farm and individual risk factors. Together, these features provide a high-resolution, One Health perspective on CCHF risk in DRC.

In southern and southeastern DRC, CCHFV seroreactivity among cattle and swine was quite high, nearly ubiquitous in some regions. This may be due to the savannah-mosaic grasslands which provide a more suitable habitat for ticks in comparison with forested regions of the country. Further models on CCHFV serosurveillance and environmental factors are needed to explore this relationship. While the observed seroreactivity indicates a high potential for CCHFV transmission among livestock and humans, only 16 *Hyalomma rufipes* were identified from ticks found on nine cattle. While *Hyalomma* ticks are the primary vector for CCHFV, the other two tick genera we identified *Amblyomma* (n = 1401, 91.4%) and *Rhipicephalus* (n = 115, 7.5%) can also transmit the virus, and these species were found in abundance^14^. Detection of the presence of CCHFV among collected ticks from seroreactive livestock can provide further understandings of CCHFV carriage among tick genera. Indeed, 87.5% of farms reported tick infestations among their herds.

We observed a statistically significant relationship between swine farms with a seroreactive animal and the rate of nose-to-nose contact between the herd and rodents. On cattle farms this relationship was not significant but around a third of farms reported rodent contact. While rodents are not considered a primary reservoir for CCHFV they may play an indirect role in the transmission chain as a host for the tick vector, an amplifier, or bridge host between questing ticks and livestock.

In general, the agricultural workers surveyed likely experience frequent low-level exposures during their job duties and lifestyles outside of work that have an additive effect over their lifetimes. Handwashing and PPE use were limited, and a majority of individuals interacted closely with sick animals. We observed a greater proportion of seropositivity among those aged 50 years or older compared with younger age groups which may indicate that there is repeated CCHFV exposure and risk across the lifecourse. Seroreactivity was not associated with any reported zoonotic exposures: visiting the forest, hunting wild animals, slaughtering or butchering wild animals, or preparing wild animals to eat. However, 80% of participants reported that they entered the forest regularly for personal reasons, indicating that environmental and zoonotic exposures are commonplace across the DRC.

Two prior DRC serosurveys reported lower—or highly variable—anti-CCHFV IgG seropositivity. In 2013, a ruminant survey in southeastern DRC detected 2/514 seropositive cattle (≈0.4%)^15^. A subsequent analysis of archived sera collected in 2017–2019 from humans and cattle, tested with the same in-house NP-ELISA used here, found 4.4% seropositivity in humans and 42.8% in cattle^16^). Differences between those estimates and ours likely reflect geography, sampling frame, host species, exposure risks, timing/seasonality, and assay performance. Longitudinal studies, coupled with entomological sampling, are needed to clarify trends in livestock infection and tick infection rates. Assay choice is also pivotal: performance varies across platforms—for example, a 2017 Ugandan study reported 12.6% seropositivity using an in-house assay versus 75% with a commercial IDVet ELISA^17^. While the in-house assay used here performed well compared to a widely used commercial kit, harmonized assays and standardized cut-offs will be essential for future CCHFV serosurveillance.

Overall, this One Health cross-sectional survey found the exposure to CCHFV over the life course of cattle to be greater than anticipated and reinforces WHO’s designation of CCHF as a priority pathogen for endemic preparedness. This survey was cross-sectional, and as such represents only a snapshot of potential CCHF risk in the DRC. Further longitudinal research can provide important understandings of CCHF risk over time. As only 16 *Hyalomma* ticks were identified, targeted research of questing ticks, rather than feeding ticks could provide important data on tick habitat in the DRC. Additional testing on collected ticks could provide further understandings of CCHFV transmission in agricultural settings.

The DRC occupies a strategically important position for zoonotic surveillance. It is home to a high density of human-livestock interaction, emerging land use change, climate-sensitive vector habitats, and limited healthcare access—all contributing factors to pathogen spillover risk. Agricultural workers are at heightened risk for zoonotic transmission and proper health reporting systems and communication regarding occupational safety should be prioritized to support CCHFV control and outbreak preparedness.

## Data Availability

All data produced in the present study are available upon reasonable request to the authors.

## References

1. Whitehouse CA. Crimean–Congo hemorrhagic fever. Antiviral Research. 2004;64(3):145–160.

2. Walker PJ, Siddell SG, Lefkowitz EJ, et al. Changes to virus taxonomy and the Statutes ratified by the International Committee on Taxonomy of Viruses (2020). Archives of Virology. 2020;165(11):2737–2748.

3. Ergönül Ö. Crimean-Congo haemorrhagic fever. The Lancet Infectious Diseases. 2006;6(4):203–214.

4. Srivastava S, Kumar S, Sharma PK, et al. Control strategies for emerging infectious diseases: Crimean-Congo hemorrhagic fever management. Health Science Reports. 2024;7(9):e70053.

5. Organization WH. Prioritizing diseases for research and development in emergency contexts. https://www.who.int/activities/prioritizing-diseases-for-research-and-development-in-emergency-contexts.

6. Hawman DW, Feldmann H. Crimean–Congo haemorrhagic fever virus. Nature Reviews Microbiology. 2023;21(7):463–477.

7. Gargili A, Estrada-Peña A, Spengler JR, Lukashev A, Nuttall PA, Bente DA. The role of ticks in the maintenance and transmission of Crimean-Congo hemorrhagic fever virus: A review of published field and laboratory studies. Antiviral Research. 2017;144:93–119.

8. Celina SS, Italiya J, Tekkara AO, Černý J. Crimean-Congo haemorrhagic fever virus in ticks, domestic, and wild animals. Frontiers in Veterinary Science. 2025;Volume 11 - 2024.

9. Atim SA, Ashraf S, Belij-Rammerstorfer S, et al. Risk factors for Crimean-Congo Haemorrhagic Fever (CCHF) virus exposure in farming communities in Uganda. Journal of Infection. 2022;85(6):693–701.

10. Ilboudo AK, Dione M, Nijhof AM, et al. Factors associated with knowledge, attitudes, and practices of mixed crop-livestock farmers on Crimean-Congo hemorrhagic fever (CCHF) and other zoonoses in Burkina Faso. One Health. 2025;20:101066.

11. Miriti M. Democratic Republic of Congo - Country Overview United States Department of Agriculture, Foreign Agricultural Service; February 26, 2025 2025.

12. Casals J. Antigenic similarity between the virus causing Crimean hemorrhagic fever and Congo virus. Proc Soc Exp Biol Med. 1969;131(1):233–236.

13. Lombe BP, Miyamoto H, Saito T, et al. Purification of Crimean-Congo hemorrhagic fever virus nucleoprotein and its utility for serological diagnosis. Sci Rep. 2021;11(1):2324.

14. Hoogstraal H. Review Article1: The Epidemiology of Tick-Borne Crimean-Congo Hemorrhagic Fever in Asia, Europe, and Africa23. Journal of Medical Entomology. 1979;15(4):307–417.

15. Sas MA, Mertens M, Kadiat JG, et al. Serosurvey for Crimean-Congo hemorrhagic fever virus infections in ruminants in Katanga province, Democratic Republic of the Congo. Ticks and Tick-borne Diseases. 2017;8(6):858–861.

16. Pongombo BLM-B, Yannick; Kashitu-Mujinga, Gracia; Mukadi, Patrick; Mampasi, Honoré; Tshilenge Mbuyi, Curé Georges; Marguerite, Manwana Pemba; Mwayakala, Noël; Zenga Bibi, Niclette; Okitale-Talunda, Patient; Pukuta, Elisabeth; Taga, Suguru; Amzati Sefu, Gaston; Kajihara, Masahiro; Nsele, Pierre Mutantu; Muyembe, Jean-Jacques; Ahuka-Mundeke, Steve; Masumu, Justin; Makiala-Mandanda, Sheila; Takada, Ayato. Seroprevalence of Crimean-Congo Hemorrhagic Fever Virus Infection in Humans and Domestic Ruminants in the Democratic Republic of the Congo: A Cross-Sectional Study. pre-print. 2025.

17. Balinandi S, von Brömssen C, Tumusiime A, et al. Serological and molecular study of Crimean-Congo Hemorrhagic Fever Virus in cattle from selected districts in Uganda. Journal of Virological Methods. 2021;290:114075.

